# Acceptance of COVID-19 vaccine among healthcare workers in Katsina state, Northwest Nigeria

**DOI:** 10.1101/2022.03.20.22272677

**Authors:** Ahmed Tijani Abubakar, Kabir Suleiman, Ahmad Suleiman Idris, Shamsuddeen Yahaya Suleiman, Umar Bello Ibrahim, Suleiman Bello Abdullahi, Ahmed Suleiman Haladu, Ahmad Ibrahim Al-Mustapha, Musa Imam Abubakar

## Abstract

High acceptance of COVID-19 vaccines is crucial to ending the COVID-19 pandemic. Healthcare workers (HCWs) are frontline responders in the fight against COVID-19, they were prioritized to receive the COVID-19 vaccine in Nigeria. This study assessed the acceptance of the COVID-19 vaccine among HCWs in Katsina State using an online structured questionnaire and predicted variables that could increase the acceptance of the vaccine among HCWs using logistic regression analysis.

A total of 793 HCWs were included in this study. Of these, 65.4% (n=519) of them were male and 36.2% (n=287) were aged between 30-39 years. Eighty percent (80%, n=638) of the HCWs have been tested for the SARS-CoV-2 out of which 10.8% (n=65) of them tested positive. The majority of the HCWs (97.3%, n=765) believed that the COVID-19 vaccine was safe and 90% (n=714) of the HCWs have received the first dose of the COVID-19 vaccine. Our findings showed that the age of the HCW, their COVID-19 testing status, and the type of health facility they work (either public or private) were the main predictors for the acceptance of the COVID-19 vaccine among HCWs in Katsina State. HCWs between the age of 30-39 years were more likely (OR:7.06; 95% CI: 2.36, 21.07; p < 0.001) to accept the vaccine than others. In the same vein, HCWs that have been tested for COVID-19 were more likely (OR:7.64; 95% CI: 3.62, 16.16; p < 0.001) to accept the vaccine than those that have not been tested. In addition, HCWs in public health facilities were more likely (OR: 2.91; 95% CI: 1.17, 6.11; p = 0.094) to accept the COVID-19 vaccine than their counterparts in private HFs.

There was a high acceptance of the COVID-19 vaccines among HCWs in Katsina State. More emphasis should be paid on adherence to non-pharmaceutical interventions and availability of vaccines for HCWs in private hospitals.

## Introduction

The coronavirus disease 2019 (COVID-19) is a major public health threat that has affected over 220 countries worldwide. As of June 15^th^ 2021, Nigeria has recorded 167, 078 COVID-19 cases and 2,117 COVID-19 related deaths out of the 176 million cases and 3.8 million deaths recorded globally [1,2].

To curb the spread of the virus, save lives, and flatten the epidemic curve, several public health measures such as social distancing, wearing of face mask, frequent hand washing, avoidance of crowded places, self-isolation, and quarantine were instituted globally. However, these measures were not sufficient [3]. Thus, humanity had the urgent need to establish herd immunity through alternative interventions such as vaccinations [4,5]. The collaborative effort of the international scientific community led to the rapid development of effective and safe vaccines against COVID–19.

Nigeria (with a population of 214 million inhabitants) prioritized the frontline healthcare workers (HCWs), elder statesmen, and the elderly persons (>60 years) as the first recipients of the 4 million doses of the AstraZeneca COVID-19 vaccine delivered to the country [6]. The uptake of the COVID–19 vaccine has however faced challenges that ranged from vaccine hesitancy and lack of trust in vaccine safety and effectiveness even among HCWs [3,7]. HCWs are crucial to Nigeria’s primary healthcare system as they provide reliable information about vaccines and vaccine-preventable diseases especially at the grassroots [8,9]. Hence, their attitude towards the COVID-19 preventive practices and their acceptance of the COVID-19 vaccine could influence the acceptance of the vaccine by the general population [10,11].

Katsina state has recorded 2,011 COVID-19 positive cases and 35 COVID-19 associated deaths since its index case on the 7^th^ of April 2020. The highest daily COVID-19 incidence in Katsina state was 70 (17^th^ December 2020) [1]. The state received 107,504 doses of the COVID-19 vaccine and has since started vaccinating its priority individuals [12]. To the best of our knowledge, this is the first study evaluating the acceptance of the COVID-19 vaccine among HCWs in Northern Nigeria. Hence, this study assessed the acceptance of the COVID-19 vaccine among HCWs in Katsina State, Nigeria.

## Materials and Methods

### Study Area

The study was conducted in Katsina State, Northwestern Nigeria. The state has 33 local government areas and a population of 5,801,584 inhabitants [13]. The state is mostly dominated by Hausa and Fulani ethnic tribes.

### Survey methodology

The survey instrument was designed to assess the acceptance of the COVID-19 vaccine by HCWs in Katsina State. The questionnaire had two sections: Section A focused on the demographics of the HCWs whereas Section B assessed the acceptance of the vaccine by HCWs. The survey instrument was validated by two independent academic reviewers to examine its content and face validity as well as its ease of administration. The questionnaire was administered as an electronic survey (via open data kit) as part of the COVID-19 operational research in Katsina State between 1^st^ and 22^nd^ May 2021. Participants were recruited by the non-probability sampling (snowballing) technique. We shared the questionnaire via social media platforms such as WhatsApp, Twitter, etc. to capture all the health facilities registered on the DHIS2 (except a few that could not be reached).

The inclusion criteria were that the participant had to be a healthcare worker. Since there were no published studies that evaluated COVID-19 vaccine acceptance in Katsina state, we assumed that 50% of the HCWs would accept the vaccine. Hence, a sample size of at least 384 respondents was needed. Study participants were selected by simple random sampling based on their willingness to participate in the study.

### Statistical analysis

The data generated from this study were analyzed using Statistical Package for Social Sciences v. 26 (Armonk, New York, USA). Descriptive statistics of the data were presented as frequencies and proportions for qualitative data as well as the mean and standard deviation for quantitative data. We conducted a univariable logistic regression analysis to evaluate the effect of the independent variables (socio-demographic variables) on the binary outcome variable (acceptance of the COVID-19 vaccine). All the significant variables at p≤ 0.05 were retained and included in the final model of the multivariable logistic regression analysis using the logit function and α at 0.25.

### Ethical approval

The ethical approval of this study was obtained from the Katsina State Ministry of Health (Reference number: MOH/KS/EHC/77/98). The objective of the study was briefly explained to the participants before a written consent form was administered to each participant. Only participants that consented were included in this survey. Participants were free to withdraw from the study at any point and no personal identifying information was obtained from any of the study participants.

## Results

A total of 793 HCWs were included in this study. Of these, 519 of them (65.4%) were male and 36.2% (n=287) were aged between 30-39 years. Most of the respondents (87.5%, n=694) were from public health facilities (Table 1). The majority of HCWs complied and practiced the laid down COVID-19 preventive measures such as regular handwashing with soap and water or using hand sanitizers. However, only around ¾ of the HCWs frequently used nose masks, maintained social distancing, and avoided crowded places (Table 2).

**Table 1.**
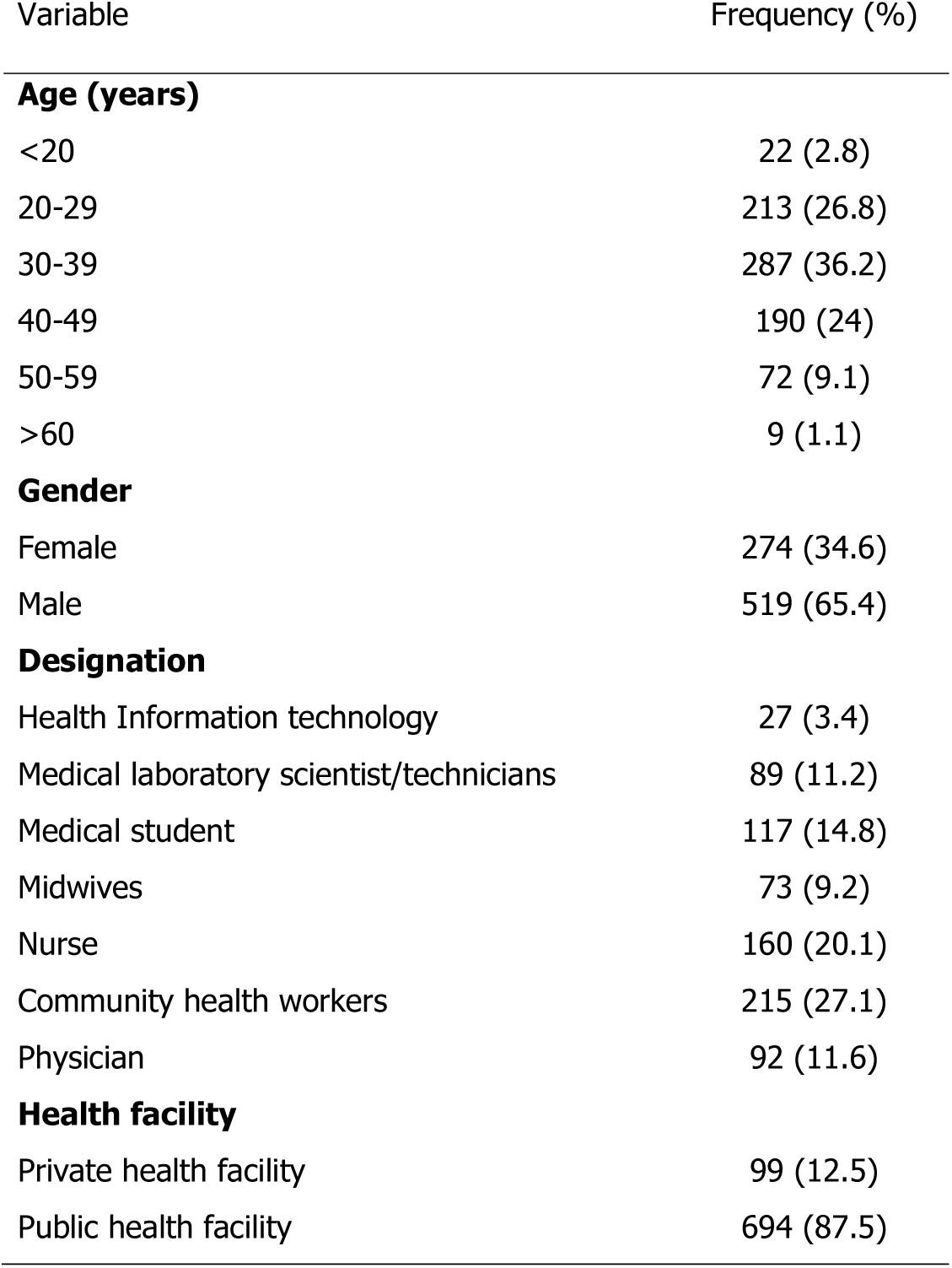
Structure of health care workers who participated

**Table 2.**
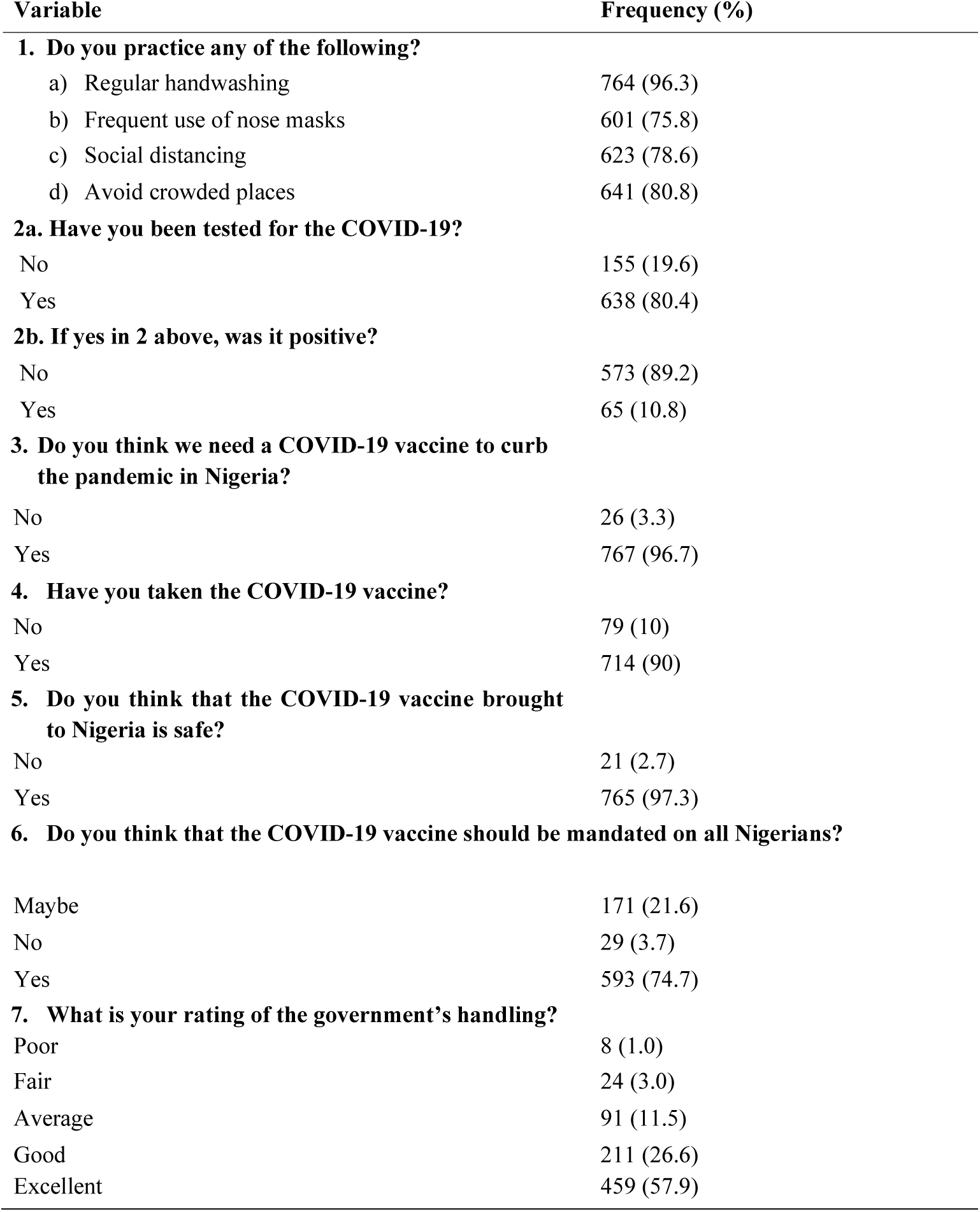
Vaccine acceptance among HCWs in Katsina State (n=793).

Eighty percent (80%, n=638) of the HCWs in both the public and private health facilities (HFs) have been tested for the COVID-19 disease either via polymerase chain reaction (PCR) or via rapid diagnostic test kits (RDT kits). Of the HCWs that have been screened for the COVID-19, 10.8% (n=65) tested positive and were managed appropriately. The majority of the HCWs (97.3%, n=765) believed that the AstraZeneca COVID-19 vacci6e received by Nigeria (from the global COVAX system) was safe and some of them (74.7%, n=573) opined that the vaccine should be mandated on all Nigerians. Ninety percent (n=714) of all HCWs who participated in this survey have received the first dose of the AstraZeneca COVID-19 vaccine. Only 4% (n=32) of all respondents were not satisfied with the government’s handling of the pandemic.

Our findings showed that the age of the HCW, their COVID-19 testing status, and the type of HF were the main predictors for the acceptance of the Covid-19 vaccine among HCWs in Katsina state. Older HCWs between the age of 30-39 were more likely (OR:7.06; 95% CI: 2.36, 21.07; p < 0.001) to accept the vaccine than others. A similar higher likelihood ratio was predicted for other groups as against young HCWs between 20-29 years. In the same vein, HCWs that have been tested for COVID-19 were more likely (OR:7.64; 95% CI: 3.62, 16.16; p < 0.001) to accept the vaccine than those that have not been tested. In addition, HCWs in public HFs were more likely (OR: 2.91; 95% CI: 1.17, 6.11; p = 0.094) to accept the COVID-19 vaccine than their counterparts in private HFs.

## Discussion

For centuries, public health interventions such as vaccination have been crucial to the prevention and control of infectious diseases [14-16]. The arrival of the COVID-19 vaccine into Nigeria marked another phase in the control of the pandemic. Our findings showed that the majority of HCWs complied and practiced the laid down COVID-19 preventive measures. HCWs complied more with regular handwashing with soap and water or using hand sanitizers as against other COVID-19 preventive practices (Table 2).

As the pandemic fatigue worsens, only around 75% of the HCWs frequently used nose masks, maintained social distancing, and avoided crowded places. This fatigue could have been further complicated by the availability of vaccines among other factors [17]. However, HCWs should strictly comply and adhere to the COVID-19 preventive practices despite the availability of COVID-19 vaccines. Moore et al,.[18] and Yu et al., [19] reported that non-pharmaceutical interventions are still needed to control the pandemic despite the global COVID-19 vaccine rollout.

Our findings showed that the prevalence of COVID-19 among HCWs in Katsina State was 10.8%. However, it remains to be ascertained whether HCWs contracted the SARS-CoV-2 via community transmission or through contact with infected patients in their respective HFs. Reports by Alasia and Maduka, [20] as well as Ogboghodo *et al*., [21] had reported a higher (15.3%) and lower (5.3%) COVID-19 prevalence rate among HCWs in Rivers and Edo states of Nigeria respectively.

Our study showed that 90% of HCWs in Katsina state have received the first dose of the vaccine. This is higher than 77% and 51% vaccine uptake reported by Maltezou *et al*., [11] among Physicians and Nursing staff respectively in Greece. The high vaccine uptake may not be unconnected to the perception of the vaccine safety by HCWs in Katsina state as 93.7% of the HCWs believed the vaccine is safe and effective. The high uptake of the vaccine by the HCW will play a great role in increasing acceptance and uptake of the vaccine by the general population since HCWs serve as trusted source of information to the general public and their advice can be a determinant of the patient’s decision to take the vaccine [3]. Similarly, HCWs highly rated the government’s handling of the pandemic as only 4% were not satisfied with the government’s handling of the pandemic. These high ratings coupled with the good perception of the COVID-19 vaccine could be responsible for the high acceptance rate [22].

Our findings showed that the main predictors of increased acceptance of the COVID-19 vaccine among HCWs in Katsina were age, COVID-19 testing status, and the type of HF. HCWs aged between 30-39 years were likely to accept the vaccine than others. This may be due to the higher percentage of this age bracket among the health sector workforce in Nigeria. A similar trend was observed by Shekhar *et al*., [23] who reported that COVID-19 vaccine acceptance increased with increasing age.

Similarly, an increased acceptance of the COVID-19 vaccine could be predicted by the testing status of HCW. Eighty percent (80%) of the study participants have been tested by either PCR or RDT kits. The willingness to get tested reflects the level of education and knowledge of the HCWs, which are additional factors that could influence vaccine acceptance by HCW. This agrees with the findings of Maltezou *et. al*., [11] and Shekhar *et al*., [23]. Vaccine acceptance was lower in a private HFs compared to public HF. This may be due to the increased risk perceptions of COVID – 19 among HCW in public HF compared to those in private HF. Perception of risk is significant for vaccine decision-making [3]. Another factor that may explain the disparity in vaccine uptake among HCW in public and private HF is the prioritization of vaccines to the frontline HCW which are majorly found in the public HF where COVID -19 cases are mostly managed, and vaccines are administered. It is however imperative to increase vaccine uptake among HCW in private HF by including them among those prioritized for vaccination and making the private HF a site of vaccination for the public. This will generally increase the number of vaccine uptake among HCW, prevent infection and reduce loss of essential workforce to infection and death [23]. By extension, it will have a positive impact on vaccine uptake among the public because of the strong association identified by Shekhar *et. al*., [23] among HCW who may intend to get vaccinated and recommend the vaccine to others.

The main strength of the study was the sample size and the timeliness of this survey. The main limitations were the sampling bias because some HFs could not access the online survey.

## Conclusion

This study reports the high acceptance of the COVID-19 vaccine among HCWs in Katsina State, Northwest Nigeria. The main predictors of an increased vaccine acceptance among HCWs were the age and the testing status of the HCW. HCWs must adhere more to COVID-19 preventive practices.

## Data Availability

The dataset generated during the survey is available on reasonable request.

## Declarations

## Acknowledgment

We acknowledge acknowledge Dr Muftau Oyewo for reviewing the manuscript.

## Author Contributions

Authors contributed equally to the conception, analysis, and writing of the manuscript.

## Data Availability

The dataset generated during the survey is available on reasonable request.

## Appendices

### Appendix 1 Tables

**Table 3.**
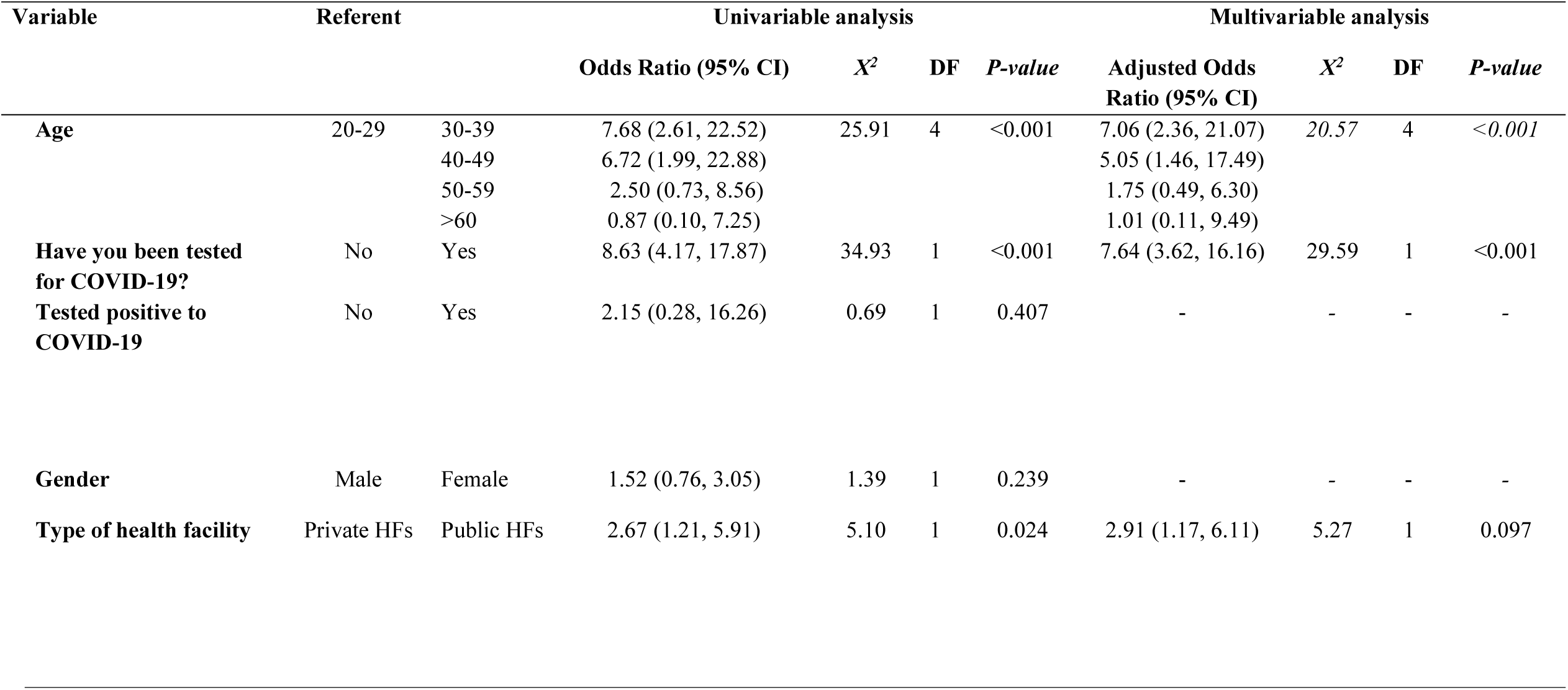
Logistic regression analysis of the factors that could predict the acceptance of the AstraZeneca COVID-19 vaccine among HCWs in Katsina State, Nigeria.

